# Clinical validity of single-gene non-invasive prenatal testing for sickle cell disease and beta thalassemia

**DOI:** 10.1101/2021.09.02.21262837

**Authors:** Erik R. Westin, David S. Tsao, Oguzhan Atay, Brian P. Landry, Patrick P. Ye, Devon Chandler-Brown, Brian Alford, Jennifer Hoskovec, Akila Subramaniam, Kevin M. Pawlik, Spencer G. Kuper, Frederick D. Goldman, Tim M. Townes, Vivien A. Sheehan

## Abstract

**Background:** Sickle cell disease (SCD) and beta thalassemia (β-thalassemia) are among the most common and severe genetically inherited disorders in the world. Although the maternal carrier status of these beta hemoglobinopathies is screened as a part of routine prenatal care in the US, the paternal carrier status is usually unavailable. Under this current screening paradigm, identification of the majority of SCD and beta thalassemia cases could therefore be delayed until newborn screening results are available. An effective reflex, non-invasive prenatal test (NIPT) that uses only maternal blood would permit prenatal detection of sickle cell disease and beta thalassemia by screening for these disorders without the need for paternal DNA. This screening would enable patients and healthcare providers to make informed decisions about diagnostic testing, and it could expand gene therapy treatment options by requesting cord blood banking at delivery. We have previously developed a single-gene NIPT platform (UNITY™) for SCD and beta thalassemia with extensive analytic validation and initial clinical validation in our prior studies.

**Objectives:** The objective of this study is to further assess the clinical validity of single-gene NIPT for SCD and beta thalassemia in a larger pregnancy population.

**Study Design:** Pregnant women who screened positive for at least one allele of a beta hemoglobinopathy were enrolled at two academic medical centers for this retrospective cohort study. Single-gene NIPT for SCD and beta thalassemia was performed in blinded fashion on maternal blood samples to determine the fetal genotype and disease status. Single-gene NIPT findings were compared with either newborn screen results or genotyping of umbilical cord blood.

**Results:** Single-gene NIPT detection of fetuses that are affected versus unaffected with SCD and beta thalassemia was 100% concordant with newborn screening follow-up data, even in challenging samples that contained a low fetal fraction (<5%) or at earlier gestational ages. Additionally, we obtained 98.5% concordance with newborn genotypes, including differentiating healthy fetal sickle cell carriers from homozygous healthy fetuses. The sensitivity of detecting fetal carrier status of beta hemoglobinopathies was 100% (90.8% to 100% CI), and the specificity was 96.4% (81.7% to 99.9 % CI).

**Conclusions:** Single-gene NIPT is a highly accurate screen for prenatal detection of SCD and beta thalassemia. The results further suggest that the maternal carrier screen with reflex single-gene NIPT workflow can significantly improve the detection rates of at-risk pregnancies from <50% to >98%.

**AJOG at a Glance:** *Why was this study conducted?:* Effective non-invasive prenatal testing (NIPT) is needed to permit safe *in utero* diagnosis of sickle cell disease (SCD) and beta thalassemia (β-thalassemia), and permit collection and banking of cord blood for future cell therapy. We have previously developed a single-gene NIPT method for SCD and β-thalassemia, with extensive analytic validation and initial clinical validation in our prior studies.

*What are the key findings?:* Our single-gene NIPT detection of SCD and β-thalassemia disease was 100% concordant with newborn screening follow-up data in 79 pregnancies at risk for beta hemoglobinopathies. Furthermore, a more stringent requirement, the detection of exact fetal genotype, was concordant in 67 out of 68 (98.5%) of pregnancies.

*What does this study add to what is already known?:* Our results validated that single-gene NIPT is an accurate and significantly improved screen for prenatal detection of SCD and β-thalassemia compared to current carrier screen workflow.

## Introduction

Hemoglobinopathies are the most common genetic disorders in the world.^1^ Beta hemoglobinopathies, including sickle cell disease (SCD) and beta thalassemia (β-thalassemia), are recessively inherited genetic disorders caused by mutations in the β-globin gene, and are often debilitating disorders that require lifelong management. SCD is the most common monogenic disorder with high prevalence among individuals originating from Sub-Saharan Africa, Middle East, South Asia, and the Mediterranean.^2^ In the United States, about 1.5% of newborns screen positive for a beta hemoglobinopathy, and over 100,000 people have SCD^3^. Among African Americans, about 1 in 12 are carriers and 1 in 365 are affected with SCD.^2,4^ Limited diversity exists among pathogenic variants related to SCD including the most common mutation, Glu6Val (HbS), and less common variants such as Glu6Lys (HbC), Glu26Lys (HbE).^5^ Mutations found in β-thalassemia are much more diverse (>200) and range from a reduction in beta globin chains (beta^+^) to mutations that lead to an absence of beta globin production (beta^0^).^6^ Compound heterozygosity of SCD and β-thalassemia variants is rare and prognoses differ depending on the underlying genotype.

Current medical guidelines, including ACOG Committee Opinion 691, recommend that all pregnancies should be carrier screened for these recessively inherited hemoglobinopathies.^7^ Screening is important for a number of reasons, including allowing parents to make informed choices, early genetic counseling, as well as optimal *in utero* and postpartum care. Additionally, prenatal testing is essential to identify fetuses for whom cord blood could be banked for future treatments. One example is gene based therapy using autologous hematopoietic stem cells (HSCs), an emergent therapeutic approach that can overcome the limitations of allogenic cell transplantation, one of which is limited donor availability.^8–12^ The first stem cell treatment for β-thalassemia has already been approved in Europe and a number of Phase I, II, and III clinical trials are underway using lentiviral or CRISPR-editing methods to treat sickle cell disease and beta thalassemia. These trials suggest that stem cell-based treatments are only a few years away from clinical use making identification of fetuses affected by hemoglobinopathies paramount, so that cord blood, as a key source of stem cells, can be banked.

However, the traditional carrier screening workflow has relatively low sensitivity for detecting high-risk fetuses due to problems associated with paternal testing.^13^ Importantly, a review of all prenatal and preconception patients seen for genetic counseling by the University of Texas Health Genetic Counseling groups found that among all women who received an abnormal carrier screening result, only 41.5% of their male partners completed screening.^14^ This suggests that at least 58.5% of affected fetuses can be missed by the current carrier screening workflow. Even if a positive paternal carrier status is obtained, the risk that the fetus is affected is at most 1 in 4. This can be an unsatisfying prior risk for patients, genetic counselors, and healthcare providers as they decide whether to obtain better information about the fetal disease status by invasive tests such as amniocentesis or chorionic villus sampling.

Therefore, there is an important unmet need for effective and accurate prenatal screening for recessive genetic disorders that does not require paternal DNA. NIPT, which analyzes cell-free fetal DNA circulating in maternal blood, has recently become standard of care for genetic aneuploidies.^15^ However, current clinical cell-free DNA (cfDNA)-based prenatal tests are limited to the detection of chromosomal abnormalities or large structural variants.^15^ Limitations of existing NIPT approaches, including the inability of these aneuploidy methods to accurately count the number of input DNA molecules, have prevented application of NIPT in hemoglobinopathies and other recessive single-gene disorders. In research contexts, digital droplet PCR-based NIPTs have been developed for monogenic, recessively inherited disorders.^16,17^ However, these tests require knowing the maternal and paternal pathogenic variants in order to design a personalized assay for each case. Furthermore, the personalized assay design can result in longer turn-around times and limited clinical laboratory throughput. To overcome these limitations, we previously developed a sequencing-based single-gene NIPT platform (UNITY™), which uses Quantitative Counting Template (QCT) molecules to simultaneously detect all common pathogenic variants for beta hemoglobinopathies in cfDNA.^18^

The objective of this study is to further assess the clinical validity of UNITY for detecting fetal beta hemoglobinopathy status (e.g. carrier or affected) in a pregnant cohort. We have previously validated this approach analytically and clinically in smaller cohorts.^18^ To further validate the utility of single-gene NIPT for SCD and beta thalassemia, herein we performed single-gene NIPT on maternal plasma obtained from a retrospective cohort of pregnant women identified as healthy carriers for SCD or beta thalassemia, or individuals affected by SCD after 10 weeks gestation, and compared the results with newborn follow-up data.

## Materials and Methods

This clinical study included pregnant women who had previously screened positive for beta hemoglobinopathy carrier status. Samples were collected from 79 subjects between October 2018 and December 2019 at the Baylor College of Medicine and University of Alabama at Birmingham with informed consent under protocols approved by institution’s internal review boards. Samples were shipped to BillionToOne’s laboratory for processing and NIPT bioinformatic analysis, with operators blinded to the fetal/newborn disease status. Ground truth outcomes were determined after birth either by chart review of state newborn screening results or by genotyping of umbilical cord blood. Study exclusion criteria were multiple gestation, unavailability of newborn ground truth, and assay failure (e.g., contamination or sequencing quality control failure).

### Sample collection, processing and NIPT analysis

Venous blood samples were collected in Streck cell-free DNA blood collection tubes, shipped to BillionToOne’s centralized laboratory, and processed for single-gene NIPT. Cell-free DNA was purified from maternal plasma, with 10 µl used to determine the fetal fraction and the remaining 35 µl of cfDNA was used for the allele fraction (AF) assay. The AF assay determines the likelihood ratio (LR) of fetal disease status based on measured diseased AF, fetal fraction, and genomic equivalents (GEs) in a relative mutation dosage (RMD) analysis.^18^ DNA purification, library preparation, bioinformatics processing, and assessing disease risk via NIPT statistical modeling were carried out as previously described.^18^

Since the paternal carrier status is unknown in these samples, an upper bound for pre-test and post-test probability of disease was calculated based on the ethnic population with the highest variant frequencies. When the mother is an unaffected carrier, the pre-test probability of fetal beta hemoglobinopathy is one fourth of the paternal carrier frequency rate. In the US, African Americans have the highest population allele frequency for beta hemoglobinopathy variants (1 in 12).^2^ Within the African American population, the highest subpopulation allele frequency is 1 in 8.^19^ The upper bound of pre-test probability of single-gene NIPT in this study is thus 1 in 32. The post-test probability is calculated from the pre-test probability and the likelihood ratio (LR) obtained from RMD analysis.^20^

The *HBB* genotypes (i.e., sickle cell carrier status) of unaffected fetuses were called based solely on a LR score without factoring in pre-test probabilities. Pre-test probabilities are less important for determining the fetus’s carrier status, since the probabilities of the fetus being a carrier versus inheriting two benign alleles are approximately equal. The likelihood ratio was calculated through RMD analysis of the benign allele, comparing the likelihood of inheriting two copies of the benign allele versus single-copy inheritance based on the allele dosage.

### Ground truth collection from umbilical cord blood

The disease status of the fetus was obtained after the child was born and underwent either universal newborn screening (NBS) or genetic analysis of umbilical cord blood. Cord blood was collected in a cord blood collection unit (Pall) via umbilical cord venipuncture. One to five microliters were removed from the collection bag under sterile conditions and lysed. Genotype was determined by the presence/absence of variant beta globin proteins in lysis solution via a lateral flow chromatographic immunological assay using HemotypeSC (Silver Lake Research) or Sickle SCAN (Biomedomics). Genotype testing was limited to HbA, HbS and HbC variants.

### Statistical analysis

Online statistics calculator MedCalc (https://www.medcalc.org/calc/diagnostic_test.php) was used to calculate specificity and sensitivity of detecting fetal HBB carrier status (homozygous unaffected versus heterozygous unaffected). Values were reported with 95% confidence interval.

## Results

### Subject demographics

Seventy-nine pregnant women identified as healthy carriers for, or affected by, a beta hemoglobinopathy at Baylor College of Medicine or UAB were enrolled in this retrospective study. Maternal beta hemoglobinopathy status included HbAS (*n* = 59), HbAC (*n* = 13), HbSS (*n* = 4), and HbCC (*n* = 1). Additionally, one patient had a whole gene HBB deletion while another was heterozygous for the beta-plus mutation HBB:c.-79A>G. The mean fetal fraction was 10.7% (range 1.1% - 23.1%). Gestational ages ranged from 16.4 weeks to collection at delivery.

### sgNIPT accurately detects fetal sickle cell disease

sgNIPT was performed on the 79 subjects. The sgNIPT analysis determines a ‘Low Risk’, ‘Decreased Risk’, ‘High Risk’, or ‘No Call’ classification. The residual risk, i.e. post-test probability, of fetal beta hemoglobinopathy is also calculated. The ground truth of whether the fetus is affected with a beta hemoglobinopathy was obtained by either chart review of the state-sponsored newborn screen or genotyping of umbilical cord blood. The sgNIPT analysis correctly identified the two newborns who had SCD, demonstrating 100% concordance (Table 1). One of the ‘High Risk’ sgNIPT cases was a sickle cell anemia affected HbSS newborn whose mother also had sickle cell anemia. The fetal fraction of sgNIPT for this case was 4.1%. The other ‘High Risk’ sgNIPT case was an HbSC newborn whose mother was heterozygous for the HbC (c.19G>A) variant. Both ‘High Risk’ sgNIPT results returned a residual risk to the fetus of >9 in 10. The vast majority of the ‘Low Risk’ calls had a fetal residual risk of 1 in 20,000; a few of the ‘Low Risk’ calls had a residual risk as high as 1 in 2,000 (Fig. 1A). There were two out of 79 ‘No Call’ results released via HBB NIPT total for a ‘No Call’ rate of 2.5%. Both of these ‘No Calls’ had a relatively low fetal fraction (1.2% and 3.8%). Newborn outcomes determined one of these newborns to be unaffected and one affected with SCD. Of note, the affected ‘No call’ was near the high end of the no call range, with a residual risk of 1 in 13, compared to a prior risk of 1 in 32 (see methods, Fig. 1A). One sample was designated as “decreased risk” because carrier screening indicated that the mother was HbAS, and the sgNIPT test detected that the fetus had inherited an HbC allele from the father. Dosage analysis on the maternal HbS allele was performed to determine that the fetus was likely an unaffected carrier with genotype HbAC, therefore lowering the residual risk from 1 in 2 to 1 in 200.

**Table 1:** Concordance between sgNIPT and outcomes for newborns affected with beta hemoglobinopathies.

|  |  | Newborn outcome |  |
| --- | --- | --- | --- |
|  |  | Unaffected | Affected |
| NIPT call | Low-risk/<br>Decreased risk | 75 | 0 |
|  | High-risk/<br>Increased risk | 0 | 2 |
|  | No-call | 1 | 1 |

**Figure 1:**
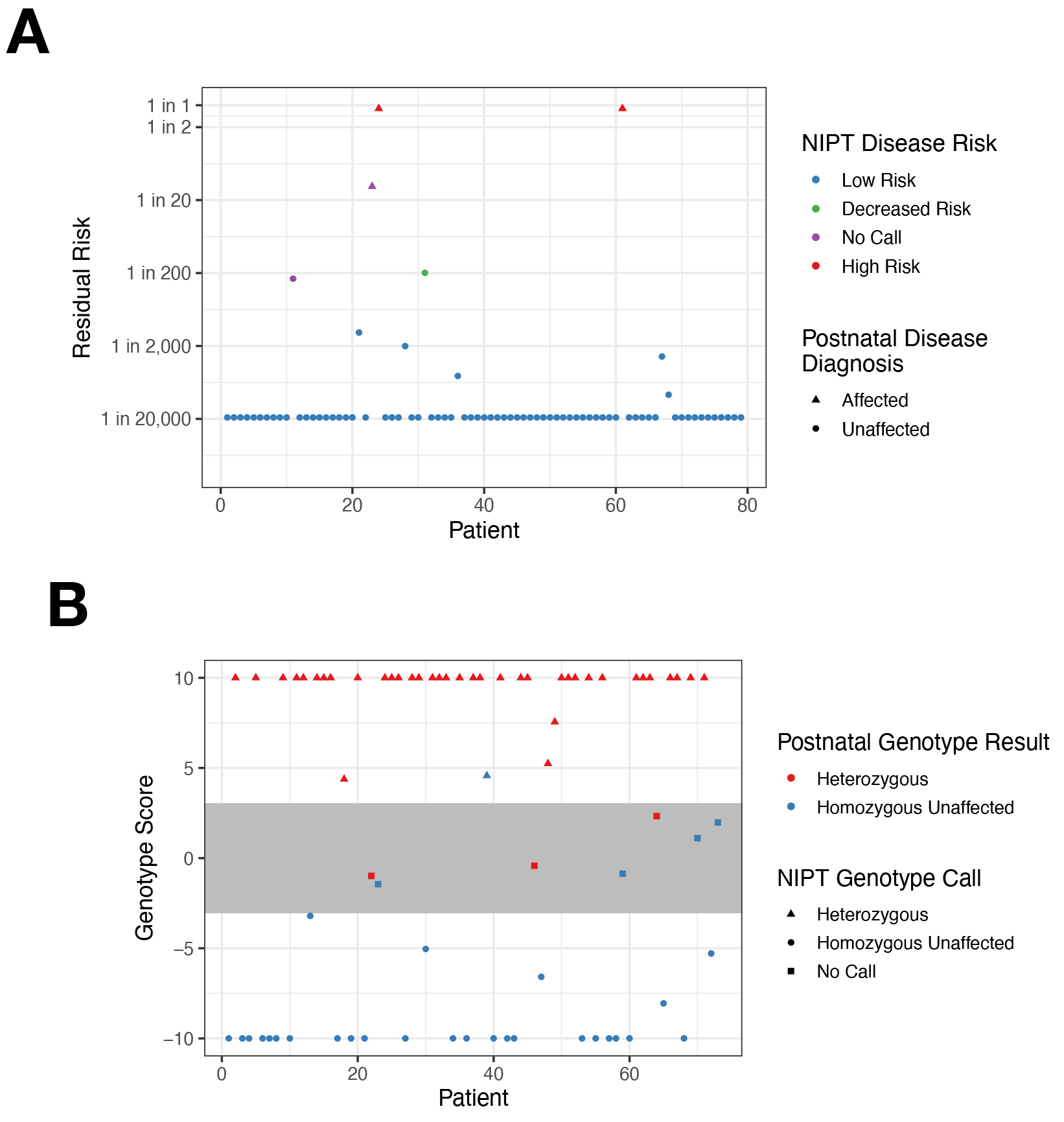
Single-gene NIPT accurately determines fetal risk and carrier status (genotype) for beta hemoglobinopathies. A. Residual risk of fetal beta hemoglobinopathy. B. NIPT differentiates between fetuses with HBB carrier (i.e. HbAC or HbAS) and homozygous unaffected (i.e. HbAA) genotypes. HBB NIPT classified a sample as genotype AA when the fetal genotype score was below -3, and as heterozygous for either HbAC or HbAS when the genotype score was above 3. Samples with scores more extreme than 10 (−10) were set to 10 (−10); scores between -3 and 3 resulted in a no call for fetal HBB carrier status. All HBB NIPT genotyping results were concordant with ground truth newborn screen, except for one sample that was determined to be AS by NIPT and AA by newborn screen.

### sgNIPT accurately determines fetal carrier status for beta hemoglobinopathies

To further demonstrate the ability of sgNIPT to detect fetal beta hemoglobinopathy variants, we next evaluated the ability of sgNIPT to predict the fetal *HBB* genotype. In general, the fetal genotype measurement is technically more challenging than an assessment of the fetal disease risk, because genotyping requires determining whether the fetus inherited 0, 1, or 2 copies of paternal and maternal variants. Classification of the fetal disease inheritance, on the other hand, only requires categorizing the inheritance pattern into either 2 copies or fewer than 2 copies.

The fetal genotype was determined for 68 of the 75 sgNIPT results. The two SCD NIPT no calls were excluded from this analysis. The two beta thalassemia carrier patients were also excluded because they were not detectable by the newborn ground truth methodology. As expected from the increased technical challenge, fetal genotyping had a higher no call rate (7 out of 75 patients) compared to fetal disease risk. Of the remaining 68 patients, sgNIPT genotype results were concordant with ground truth newborn genotype results in all but one sample (Table 2). The strength of the statistical evidence for inheriting 0, 1, or 2 copies of each variant is summarized by a fetal genotype score (Figure 1B). Samples with a genotype score above a threshold are classified as heterozygous and below are homozygous; scores close to 0 indicate a relative lack of evidence in either direction. To avoid returning uncertain results, scores that are in the range [-3, +3] were classified as no calls, consistent with evidence thresholds used previously.^17,18^ In the only discordant sample, which was close to the no call cutoff, sgNIPT analysis returned a false positive fetal genotype of HbAS when the newborn genotype was HbAA (Figure 1B). Overall, sgNIPT demonstrated a 100% (90.8% - 100%, 95% CI) sensitivity and 96.4% (81.7% - 99.9%, 95% CI) specificity for detecting whether the fetus is a carrier of a beta hemoglobinopathy.

**Table 2:** Concordance between sgNIPT and outcomes for newborn HBB genotypes.

|  |  | Newborn outcome |  |  |  |  |
| --- | --- | --- | --- | --- | --- | --- |
|  |  | Normal | Traits |  | Disease |  |
|  |  | AA | AS | AC | SC | SS |
| NIPT call | AA | 27 | 0 | 0 | 0 | 0 |
|  | AS | 1 | 31 | 0 | 0 | 0 |
|  | AC | 0 | 0 | 7 | 0 | 0 |
|  | SC | 0 | 0 | 0 | 1 | 0 |
|  | SS | 0 | 0 | 0 | 0 | 1 |

## Discussion/Comments

### 1. Principal Findings

Our NIPT approach^18^ for beta hemoglobinopathies was validated in the retrospective cohort study reported here. Out of the 79 carrier or affected pregnancies screened, sgNIPT detected two fetuses that were high risk for a beta hemoglobinopathy; this was 100% concordant with the newborn results. Crucially, sgNIPT was able to evaluate the fetal risk even though paternal genotype data was unavailable for all of these subjects. Our ability to discern fetal sickle cell carrier status further validated the accuracy of our approach, since this is a more technically challenging task than determining the fetal SCD status. Overall, there were 28 non-carriers and 38 sickle cell carriers. The sensitivity of detecting fetal carriers of beta hemoglobinopathies 100% (90.8% to 100%, 95% CI), and the specificity was 96.4% (81.7% to 99.9 % CI).

### 2. Clinical Implications

Our data suggest that our sgNIPT can be used to effectively and accurately screen for beta hemoglobinopathies in all pregnancies. Adaptation of this noninvasive, accurate, and affordable single-gene NIPT in the clinics should lead to increased numbers of parents being informed of SCD or beta thalassemia status of their child before birth, even in the 50% of instances when paternal DNA is unavailable.^14^ Accurate fetal risk assessment can provide information desired by many patients to aid in making informed decisions regarding prenatal diagnostic testing. Additionally, the personalized information can help prepare parents for delivery and enable cord blood banking of affected newborns, thereby preserving a valuable source of hematopoietic stem cells (HSCs) for rapidly emerging treatment options based on gene therapy. Compared to the current carrier screening workflow, where <42% of affected fetuses may be detected,^14^ single-gene NIPT had a 100% sensitivity for detecting fetal hemoglobinopathy in this study.

### 3. Research Implications

The NIPT for SCD and beta thalassemia that has been clinically validated in this study could enable development of effective cures that can be applied to neonates at early stages of life when gene therapies are typically more effective. As a byproduct of oxygen exchange *in utero*, infants are temporarily protected from the effects of SCD and β-thalassemia mutations for approximately one year while fetal hemoglobin (gamma-hemoglobin) expression remains elevated and adult hemoglobin (beta hemoglobin, location of the mutations) is suppressed.^21^ After one year, morbidities associated with these diseases begin to accrue and most are irreversible even upon successful stem cell transplantation. NIPT can identify fetuses for whom cord blood should be banked for future research and development of treatments based on gene editing of autologous stem cells. It is critical that this readily available source of HSCs not be discarded, as the HSCs of patients with SCD will be invaluable as a potential curative modality, especially early in life. Accurate NIPT for SCD and β-thalassemia would allow parents to request cord blood banking of newborns that have these conditions, thereby preserving a valuable source of HSCs for future research and therapies.

### 4. Strengths and Limitations

In the present study, we demonstrated the clinical validity of our sgNIPT for SCD and beta thalassemia by analysis of a retrospective cohort of pregnancies and newborn follow-up data. Until our approach, NIPT for single-gene recessive disorders had remained technically challenging. We have met this challenge with a highly accurate (even in samples with less than 5% fetal fraction of cfDNA) and resource-efficient test. Our sgNIPT detection of two fetuses with beta hemoglobinopathies was 100% concordant with newborn results. Even though there were only a small number of SCD affected fetuses, our test also correctly determined the sickle cell carrier status of 67 out of 68 fetuses (98.5% concordant with newborn results). This level of accuracy is more suited to clinical use, compared to a previous digital PCR-based single-gene NIPT that reported genotype concordance of only about 80%.^16^ A second approach that relies on haplotyping has also been proposed.^22^ While accurate, this method requires expensive whole-genome sequencing of both parents and a highly technical assay resulting in high costs and long turn-around-time, both of which preclude it from routine clinical use. In contrast, the sgNIPT assay presented here does not require paternal DNA, is significantly more cost effective, and can have faster turnaround time.

Based on the promising results of this retrospective clinical study, subsequent research is needed to enroll large number of patients in a prospective study to evaluate use of combined carrier screening and reflex sgNIPT for improving clinical care. In the current study, we enrolled pregnant patients identified as healthy carriers or affected mothers for beta hemoglobinopathies. In a future prospective study, establishing the study cohort by enrolling all patients who are undergoing first trimester prenatal screening would be more representative of the intended clinical use case.

### 5. Conclusions

Our NIPT analysis in a retrospective cohort of 79 pregnant women identified as healthy carriers or affected mothers for SCD or beta thalassemia showed that single-gene NIPT is a highly accurate approach for prenatal detection of these diseases. Combined with the unique workflow of reflex single-gene NIPT for carrier mothers without the need for a paternal sample, this screen allows for an efficient and accurate fetal risk assessment for beta hemoglobinopathies in pregnant women.

## Data Availability

Please inquire about data availability

## Highlights

- Non-invasive prenatal testing for sickle cell disease and β-thalassemia is accurate
- Non-invasive prenatal testing was validated in retrospective cohort studies
- Adaptation of non-invasive prenatal testing can improve clinical care for patients

## Acknowledgements

Research reported in this publication was supported by the National Heart, Lung, and Blood Institute of the National Institutes of Health under award number R43HL144322. In the study, 10% was funded by federal sources, and 90% was funded by BillionToOne. The content is solely the responsibility of the authors and does not necessarily represent the official views of the National Institutes of Health. We thank Dr. Rong Mao and Dr. Jacqueline Carozza for providing editorial support.

## Notes

**Disclosure Statement:** DST, OA, BL, PPY, DCB, BA and JH are employees of BillionToOne and hold stock or options to hold stock in the company. VAS received a grant from BillionToOne directly and as a subcontractor from NIH. ERW, AS, KAP, SGJ, FDG, and TMT report no conflict of interest.

**Source of Financial Support:** Research reported in this publication was supported by the National Heart, Lung, and Blood Institute (BillionToOne, R43HL144322) and by the National Institute of Diabetes and Digestive and Kidney Diseases (VAS, 1K08DK110448-01). Funding was also provided by BillionToOne, Inc. The content is solely the responsibility of the authors and does not necessarily represent the official views of the National Institutes of Health.

### Competing Interest Statement

DST, OA, BL, PPY, DCB, BA and JH are employees of BillionToOne and hold stock or options to hold stock in the company. VAS received a grant from BillionToOne directly and as a subcontractor from NIH. ERW, AS, KAP, SGJ, FDG, and TMT report no conflict of interest.

### Funding Statement

Research reported in this publication was supported by the National Heart, Lung, and Blood Institute (BillionToOne, R43HL144322) and by the National Institute of Diabetes and Digestive and Kidney Diseases (VAS, 1K08DK110448-01). Funding was also provided by BillionToOne, Inc. The content is solely the responsibility of the authors and does not necessarily represent the official views of the National Institutes of Health.

### Author Declarations

Oversight approval was made by UAB and BCM committees. Websites can be found here, UAB: https://www.uab.edu/research/home/irb and BCM https://www.bcm.edu/healthcare/clinical-trials/institutional-review-board. These protocols were IRB-38457, approved 9/14/2011 (Baylor) and IRB-160309003, approved 4/04/2016 (UAB.)IRB for research Samples were collected from 79 subjects between October 2018 and December 2019 at the Baylor College of Medicine and University of Alabama at Birmingham with informed consent under protocols approved by institution's internal review boards.

